# Synthetic Cathinones: ‘Bath Salts’ or ‘Flakka’ Poisonings and Use in the United States 2021 to 2023

**DOI:** 10.64898/2026.03.27.26349556

**Authors:** Orrin D. Ware

## Abstract

Synthetic cathinones, colloquially called bath salts or flakka, are a group of psychoactive substances used recreationally, including mephedrone and eutylone. Studies examining the prevalence of bath salt use among select samples in the United States have found that approximately 1% of nightclub attendees in New York, high school seniors, and college students have used them. The purpose of this study was to examine the national prevalence of lifetime and past-12-month use of bath salts among a nationally representative sample of persons in the United States from 2021 to 2023. This study also examined nationwide poison center data to identify the number of poisonings from 2021 to 2023 in which bath salt use was intentional and not necessarily an adulterant in another illicitly obtained recreational substance. This study identified the prevalence of lifetime bath salt use among a nationally representative sample of persons 12 years and older in the U.S. to be 0.2% (n = 670,611) in 2021, 0.3% (n = 838,941) in 2022, and 0.3% (n = 836,128) in 2023. The national prevalence of past-12-month bath salt use was 0.0% (n = 111,039) in 2021, 0.1% (n = 167,815) in 2022, and 0.1% (n = 152,276) in 2023. From 2021 to 2023, there were 148 cases in which bath salt use was intentional and involved in a reported poisoning to one of the 55 poison centers in the U.S. Future studies are needed to examine risk factors associated with bath salt-related poisonings.

## INTRODUCTION

Synthetic cathinones are used recreationally and produce a stimulating psychoactive effect, and are colloquially known by many names, including bath salts and flakka (other names include White Magic and Red Devil).^1, 2^ There are hundreds of synthetic cathinone derivatives on the clandestine drug market, including mephedrone, eutylone, pentylone, and 3,4-Methylenedioxypyrovalerone.^1, 3, 4^ New derivatives are often introduced to the drug market to replace those that are made illegal. Bath salts are prominent in the drug market, with eutylone, for example, being among the top ten most frequently identified drugs by law enforcement in the United States during the first half of 2021.^5^

The psychoactive effects of bath salts share effects that are similar to amphetamine, providing euphoria and altering brain reward systems.^1, 3, 4, 6^ Depending on the derivative, some pre-clinical models have found stronger reinforcing potency and reinforcing efficacy, such as 3,4-Methylenedioxypyrovalerone, compared to cocaine.^1, 7, 8^ The adverse effects of bath salts can include agitation, mania, hallucinations, and nausea.^1, 4, 9-12^ Prominent pulmonary issues, including cardiac arrest and tachycardia, can also emerge from bath salt-related poisoning.^4, 11, 13-16^ Fatal intoxications in which bath salts are involved have also been identified.^5, 17^ For example, eutylone was involved in 343 deaths in 2020 based on data from the Centers for Disease Control and Prevention.^5^ Further, data suggest that bath salts are occasionally sold as 3,4-Methylenedioxymethamphetamine (MDMA) with these substances typically being co-involved with overdoses related to illicitly manufactured fentanyl.^5^ Therefore, as an adulterant, some individuals may inadvertently ingest bath salts while seeking to obtain other substances on the illicit drug market.

Studies examining the prevalence of bath salt use among select samples in the United States have found that approximately 1% of high school seniors,^18, 19^ college students,^20^ and New York nightclub attendees ^21^ have used bath salts. However, there is limited literature on the national prevalence of lifetime and past-year bath salt use in the United States. Additionally, data are lacking on the estimated rate of past-year involvement of bath salts in reported poisonings among people who used bath salts in the past year. This study leverages nationally representative data on bath salt use and reported poisonings in the United States from 2021 to 2023. Findings from this study build upon prior literature by adding an update to the prevalence of bath salt use and providing annual rates of bath salt poisonings during the analytic period.

## MATERIALS AND METHODS

### Ethical Considerations

The University of North Carolina at Chapel Hill is the Institutional Review Board of record, and this study is not human subjects research. Only publicly available data were used, and therefore, informed consent is not applicable.

### Data Source and Sample Selection

#### National Survey on Drug Use and Health (NSDUH)

As one of two datasets used for this study, the National Survey on Drug Use and Health (NSDUH) is an annual nationally representative health survey of U.S. households, and the 2021 to 2023 file contains a total of 173,808 unweighted cases.^22^ Health topics such as prevalence of suicidal ideation, substance use, and receipt of treatment are captured from civilian noninstitutionalized persons aged 12 years and older in the NSDUH. By utilizing sample weights, the data may be used to provide national estimates of health topics captured in the survey. A question in the dataset assesses whether participants have ever used bath salts, “The next question is about synthetic stimulants that people use to get high, also called “bath salts” or flakka. Have you ever, even once, used these synthetic stimulants?” This question was used to select the study sample, with a binary recoded variable in the dataset indicating whether the respondent reported “Never used” or “Ever used” bath salts. Respondents flagged in the dataset as “Ever used” bath salts were included in the analytic sample. This resulted in an unweighted sample of 534 respondents.

#### Annual Report of the National Poison Data System® (NPDS) from America’s Poison Centers®

Each year, data from all 55 of America’s Poison Centers are published in an annual report describing poison exposures for a specific year. This study used data from the 2021 to 2023 Annual Report of the National Poison Data System® (NPDS) from America’s Poison Centers®.^23-25^ Case data from all 55 poison centers are added to the data warehouse called the NPDS. The annual report includes count data about specific substances, such as the number of times a substance was mentioned in a case including substance combinations in which multiple drugs were involved, the number of times a substance was the only one mentioned in a case, and the number of times the substance was used intentionally (such as self-harm, misuse, and intentional abuse). Count data for “Synthetic Cathinones, Analogs and Precursors” were extracted from the poison center annual reports for this study, specifically total case mentions, total number of single substance case mentions, and total number of cases in which use was intentional.

### Variables

Variables extracted from the NSDUH for this study include: age, population density, race and ethnicity, sex, year, and last use of bath salts. Age is comprised of six categories, identifying the respondent’s age: 12-17 years and 65 or older. Population density describes whether the respondent lives in a large metropolitan, small metropolitan, or nonmetropolitan area. Race and ethnicity includes seven categories, such as Non-Hispanic Asian and Hispanic. Year describes whether the survey data was captured in 2021, 2022, or 2023. Last use of bath salts includes three categories: [a] within the past 30 days, [b] More than 30 days ago but within the past 12 months, and [c] more than 12 months ago.

### Analysis

Statistical Product and Service Solutions (SPSS) Version 31.0 was used to apply sample weights and run univariable statistics with the NSDUH data. Lifetime and past-12-month prevalence of bath salt use were examined in the total population. After identifying the prevalence in the general population, the sample of individuals who had ever reported bath salt use was selected to further characterize these individuals. After describing the sample with lifetime use of bath salt, the sample of individuals with past 12-month bath salt use was selected and described. The ‘epiR’^26^ package was used in R Version 4.5.2 “[Not] Part in a Rumble”^27^ to identify an estimate for the rate of reported bath salt poisonings per 10,000 with the 95% Confidence Interval. To determine the rate, cases were defined as the number of poisonings in which bath salt use was intentional, and the population was defined as the number of weighted cases of individuals who reported using bath salts in the past 12 months. These rates were calculated for each year of the analytic period, 2021 to 2023.

## RESULTS

### Characterizing Individuals Who Have Ever Used Bath Salts

From 2021 to 2023, 0.3% (N = 2,345,680) of persons 12 years and older in the U.S. ever used bath salts. The national prevalence was 0.2% (n = 670,611) in 2021, 0.3% (n = 838,941) in 2022, and 0.3% (n = 836,128) in 2023. Table 1 presents aggregated data for all three years and annual data for the sample of N = 2,345,680 individuals who have ever used bath salts. Across the aggregate and yearly data, respondents primarily reside in large metropolitan areas, with the percentages ranging from 39.1% to 45.5% depending on the specific year. The aggregate data reflected that approximately two in three of the individuals who ever used bath salts were males, whereas one in three were females. Non-Hispanic White individuals account for approximately one in four persons who have ever used bath salts. Lastly, four in five persons who have ever used bath salts have not used it in the past 12 months.

**Table 1.** Aggregate and Annual Characteristics of People who Ever Used Bath Salts Recreationally from 2021 to 2023; N = 2,345,680.

|  |  | Total: 2021 to 2023<br>N = 2,345,680 |  | 2021<br>N = 670,611 |  | 2022<br>N = 838,941 |  | 2023<br>N = 836,128 |  |
| --- | --- | --- | --- | --- | --- | --- | --- | --- | --- |
|  |  | Column N |  | Column N |  | Column N |  | Column N |  |
|  |  | Count | % | Count | % | Count | % | Count | % |
| Year of Data Collection | 2021 | 670,611 | 28.6% | 670,611 | 100.0% | 0 | 0.0% | 0 | 0.0% |
|  | 2022 | 838,941 | 35.8% | 0 | 0.0% | 838,941 | 100.0% | 0 | 0.0% |
|  | 2023 | 836,128 | 35.6% | 0 | 0.0% | 0 | 0.0% | 836,128 | 100.0% |
| Population Density | Large Metro | 1,009,698 | 43.0% | 305,436 | 45.5% | 327,988 | 39.1% | 376,273 | 45.0% |
|  | Small Metro | 726,964 | 31.0% | 186,051 | 27.7% | 265,855 | 31.7% | 275,059 | 32.9% |
|  | Nonmetro | 609,018 | 26.0% | 179,124 | 26.7% | 245,098 | 29.2% | 184,796 | 22.1% |
| Age | 12-17 Years Old | 102,060 | 4.4% | 21,850 | 3.3% | 42,636 | 5.1% | 37,574 | 4.5% |
|  | 18-25 Years Old | 217,609 | 9.3% | 69,369 | 10.3% | 78,671 | 9.4% | 69,569 | 8.3% |
|  | 26-34 Years Old | 716,148 | 30.5% | 204,271 | 30.5% | 340,313 | 40.6% | 171,564 | 20.5% |
|  | 35-49 Years Old | 945,371 | 40.3% | 171,160 | 25.5% | 267,768 | 31.9% | 506,444 | 60.6% |
|  | 50-64 Years Old | 262,784 | 11.2% | 129,920 | 19.4% | 88,466 | 10.5% | 44,398 | 5.3% |
|  | 65 or Older | 101,707 | 4.3% | 74,040 | 11.0% | 21,088 | 2.5% | 6,579 | 0.8% |
| Race/Ethnicity | NH White | 1,805,448 | 77.0% | 573,649 | 85.5% | 616,871 | 73.5% | 614,927 | 73.5% |
|  | NH Black/African American | 176,540 | 7.5% | 29,963 | 4.5% | 98,564 | 11.7% | 48,013 | 5.7% |
|  | NH Native American/Alaska Native | 16,323 | 0.7% | 4,618 | 0.7% | 5,424 | 0.6% | 6,281 | 0.8% |
|  | NH Native Hawaiian/Other Pacific Islander | 51,902 | 2.2% | 7,957 | 1.2% | 6,667 | 0.8% | 37,278 | 4.5% |
|  | NH Asian | 25,924 | 1.1% | 24 | 0.0% | 2,971 | 0.4% | 22,930 | 2.7% |
|  | NH more than one race | 96,459 | 4.1% | 31,953 | 4.8% | 51,998 | 6.2% | 12,507 | 1.5% |
|  | Hispanic | 173,085 | 7.4% | 22,448 | 3.3% | 56,445 | 6.7% | 94,192 | 11.3% |
| Sex | Male | 1,480,519 | 63.1% | 434,085 | 64.7% | 513,751 | 61.2% | 532,683 | 63.7% |
|  | Female | 865,161 | 36.9% | 236,526 | 35.3% | 325,190 | 38.8% | 303,445 | 36.3% |
| Last use of Bath Salts | Within the past 30 days | 305,189 | 13.0% | 87,826 | 13.1% | 116,857 | 13.9% | 100,506 | 12.0% |
|  | More than 30 days ago but within the past 12 months | 125,941 | 5.4% | 23,213 | 3.5% | 50,958 | 6.1% | 51,770 | 6.2% |
|  | More than 12 months ago | 1,914,550 | 81.6% | 559,571 | 83.4% | 671,126 | 80.0% | 683,852 | 81.8% |

NH: Non-Hispanic

### Characterizing Individuals Who Used Bath Salts in the Past 12 Months

From 2021 to 2023, 0.1% (n = 431,130) of persons 12 years and older in the U.S. used bath salts in the past 12 months. The national prevalence was 0.0% (n = 111,039) in 2021, 0.1% (n = 167,815) in 2022, and 0.1% (n = 152,276) in 2023. Table 2 presents aggregated data for all three years and annual data for the sample of N = 431,130 individuals who used bath salts in the past 12 months. Non-Hispanic White individuals account for 44.4% to 55.4% of individuals who used bath salts in the past 12 months for each year from 2021 to 2023. Among individuals who used bath salts in the past 12 months, 66.0%-79.1% used them in the past 30 days and 20.9% to 34.0% used them more than 30 days ago.

**Table 2.**
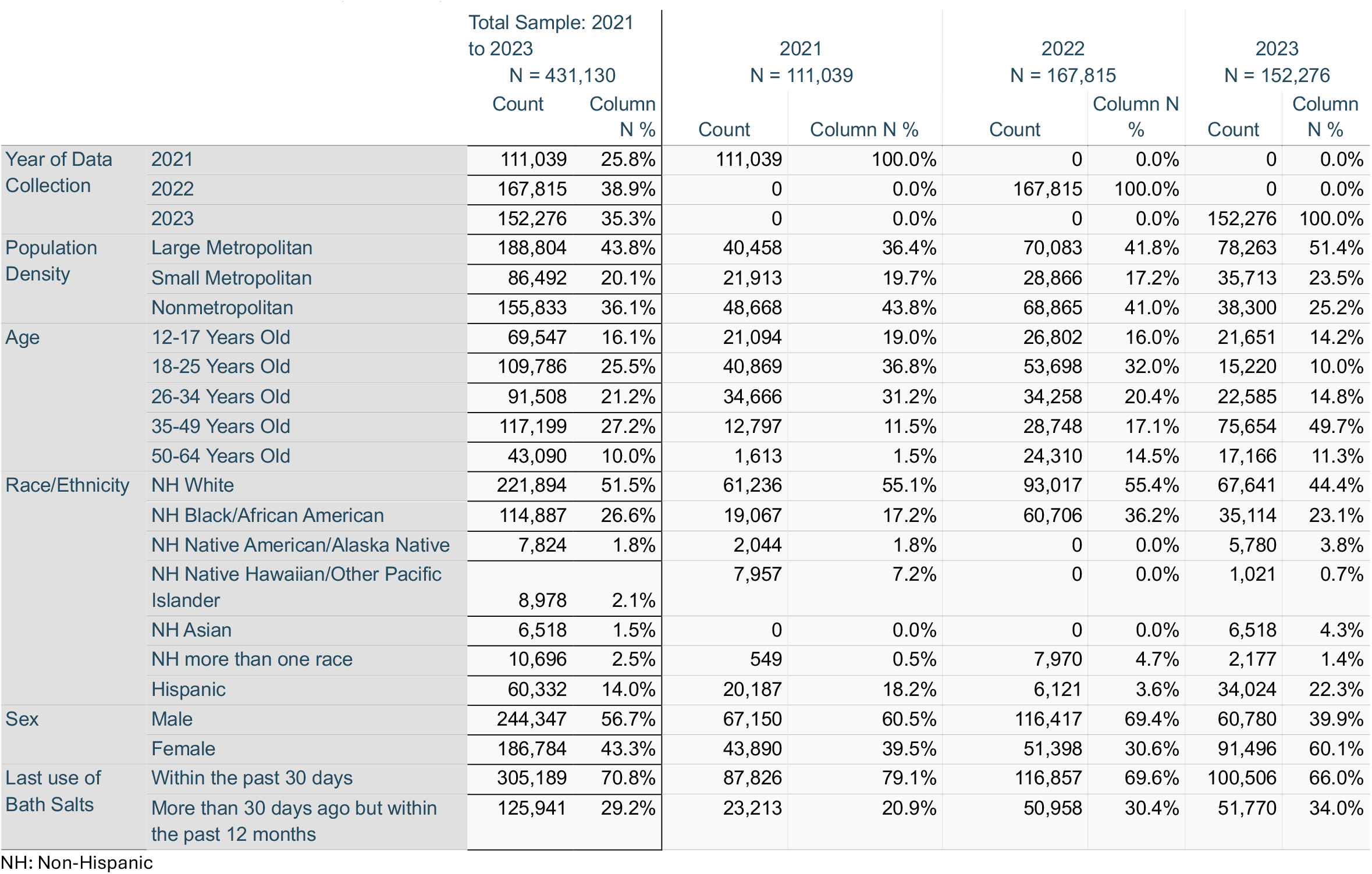
Aggregate and Annual Characteristics of People who Used Bath Salts in the Past Year from 2021 to 2023; N = 431,130.

### Estimated Rate of Bath Salt Poisonings

Table 3 presents annual bath salt poisoning counts from 2021 to 2023. Using the annual counts of intentional exposures in which bath salts were used intentionally and the annual counts of persons who used bath salts in the past 12 months, the estimated rate of bath salt poisoning was identified. In 2021, the rate of bath salt poisoning among people who used bath salts in that year was 6.2 per 10,000 (95% Confidence Interval: 4.8; 7.9). In 2022, the rate of bath salt poisoning among people who used bath salts in that year was 2.4 per 10,000 (95% Confidence Interval: 1.7; 3.2). In 2023, the rate of bath salt poisoning among people who used bath salts in that year was 2.6 per 10,000 (95% Confidence Interval: 1.8; 3.5).

**Table 3.**
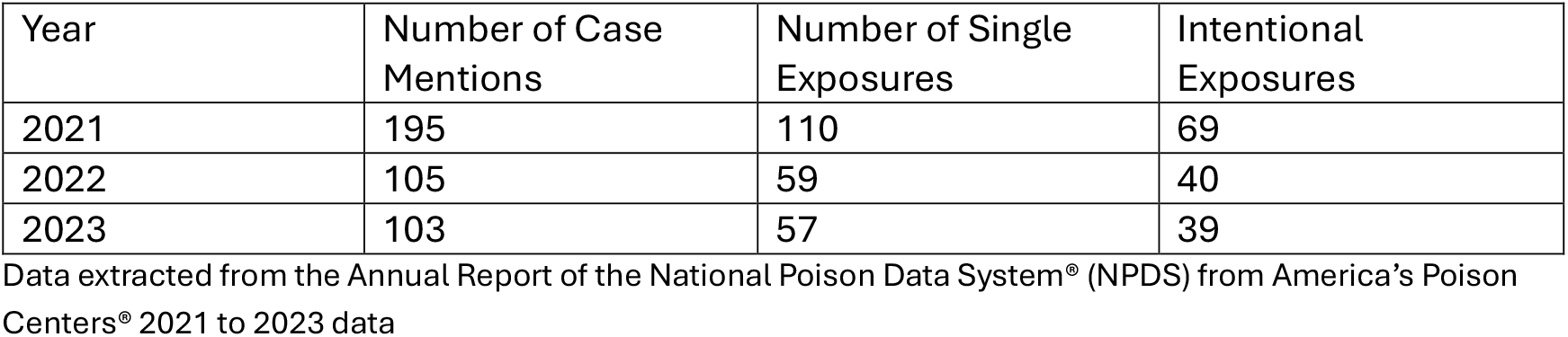
Annual Synthetic Cathinones Poison Data from 2021 to 2023.

## DISCUSSION

This study identified the prevalence of lifetime bath salt use among a nationally representative sample of persons 12 years and older in the U.S. Further, this study provides estimated rates of bath salt poisonings among persons who intentionally used bath salts. For each year of data included in this study (2021-2023), the prevalence of reported lifetime use of bath salts was low, ranging from 0.2% to 0.3%. Other studies have identified the low prevalence of using bath salts among specific samples. For example, nationally representative studies of high school seniors found that approximately 1% of the sample reported using bath salts.^18, 19^ A similar prevalence was observed among nightclub attendees in New York, with 1.1% testing positive for bath salts.^21^ A self-report survey of college students in the United States also identified approximately 1% of the sample as having ever used bath salts.^20^

An anonymous online survey that collected data from 113 individuals who reported using bath salts found that the participants were mostly males, Non-Hispanic White, and between the ages of 18 and 24 years old.^6^ Furthermore, 26% of the online sample reported no bath salt use in the past 12 months.^6^ Similarly, this current study found that the majority of individuals who had used bath salts at least once in their lifetime and in the past 12 months were Non-Hispanic White. However, among respondents in this current study who have ever used bath salts, approximately four in five have not used bath salts in the past 12 months.

From 2021 to 2023, this study identified 148 cases in which bath salt use was intentional and involved in a reported poisoning. This study focused on poisonings in which the use was intentional because some individuals may have used bath salts and been unaware, as they are often used as an adulterant, such as being sold as MDMA.^5, 21^ The estimated rates per 10,000 of experiencing a bath salt-related poisoning among persons who used bath salts in the past year were 6.2 in 2021, 2.4 in 2022, and 2.6 in 2023. The route of administration can impact the potential risk and severity of poisoning. The intranasal route of administration is commonly used for bath salts, with oral consumption of pills and tablets also being consumed.^4, 6, 28^ However, some of the bath-salt-involved poisonings likely included cases in which there was a combination of other substances, such as fentanyl or methamphetamine, also being involved.

Overdose death data involving bath salts suggest that these overdoses often involve illicitly manufactured fentanyl.^5^ Further, opioids and stimulant drug classes are often co-used in what is colloquially described as speedball. Bath salts are usually cheaper than other stimulants, such as illicit amphetamine and cocaine, presenting as an economic motivation for use.^12, 29-31^ Another consideration regarding bath salts is the variability in the psychoactive effect based on the specific drug that falls under the category of bath salts, such as mephedrone, eutylone, pentylone, and 3,4-Methylenedioxypyrovalerone, producing different psychoactive effects.^4^

### Limitations

Although this study provides the prevalence of bath salt use among the U.S. population and estimates of bath salt-involved poisonings among individuals who endorsed past-year use from 2021 to 2023, findings must be considered alongside study limitations. While the NSDUH is a nationally representative sample, it is limited by self-selection bias, self-report bias, and other biases present in national self-report studies. Individuals may have more than one poisoning, which may impact these estimates. Another limitation is that some individuals did not respond to the question on bath salt use in the NSDUH. The poison center data included in the study consist only of cases reported to poison centers; however, other poisonings may have occurred that were not necessarily reported. The count data on cases in which bath salt use was intentional also include cases in which self-harm was the intended purpose of use. Another limitation is combining weighted health survey data and real-world poison center data, as one is a nationally representative sample whereas the other is actual real-world count data. However, this provides the best estimate of nationwide self-reported use of bath salts.

## CONSLUSION

Synthetic cathinones, colloquially called bath salts or flakka, are a group of psychoactive substances used recreationally, including mephedrone and eutylone. This study identified the prevalence of lifetime bath salt use among a nationally representative sample of persons 12 years and older in the U.S. to be 0.2% (n = 670,611) in 2021, 0.3% (n = 838,941) in 2022, and 0.3% (n = 836,128) in 2023. The national prevalence of past-12-month bath salt use was 0.0% (n = 111,039) in 2021, 0.1% (n = 167,815) in 2022, and 0.1% (n = 152,276) in 2023. From 2021 to 2023, there were 148 cases in which bath salt use was intentional and involved in a reported poisoning to one of the 55 poison centers in the U.S. The estimated rates per 10,000 of experiencing a bath salt-related poisoning among persons who used bath salts in the past year were 6.2 in 2021, 2.4 in 2022, and 2.6 in 2023. This study provides the prevalence of bath salt use in the U.S. and estimates of bath salt-related poisoning. Future studies are needed to examine risk factors associated with bath salt-related poisonings.

## Declaration of Conflicting Interest

There are no conflicts to declare.

## Funding statement

There is no funding to report.

## Data Availability Statement

Datasets used in this study are located online: [1] https://www.samhsa.gov/data/data-we-collect/nsduh-national-survey-drug-use-and-health/national-releases [2] https://doi.org/10.1080/15563650.2022.2132768 [3] https://doi.org/10.1080/15563650.2023.2268981 [4] https://doi.org/10.1080/15563650.2024.2412423.

## Notes

### Competing Interest Statement

The authors have declared no competing interest.

